# Evaluating the use of GPT-3.5-turbo to provide clinical recommendations in the Emergency Department

**DOI:** 10.1101/2023.10.19.23297276

**Authors:** Christopher Y.K. Williams, Brenda Y. Miao, Atul J. Butte

## Abstract

The release of GPT-3.5-turbo (ChatGPT) and other large language models (LLMs) has the potential to transform healthcare. However, existing research evaluating LLM performance on real-world clinical notes is limited. Here, we conduct a highly-powered study to determine whether GPT-3.5-turbo can provide clinical recommendations for three tasks (admission status, radiological investigation(s) request status, and antibiotic prescription status) using clinical notes from the Emergency Department. We randomly select 10,000 Emergency Department visits to evaluate the accuracy of zero-shot, GPT-3.5-turbo-generated clinical recommendations across four different prompting strategies. We find that GPT-3.5-turbo performs poorly compared to a resident physician, with accuracy scores 24% lower on average. GPT-3.5-turbo tended to be overly cautious in its recommendations, with high sensitivity at the cost of specificity. Our findings demonstrate that, while early evaluations of the clinical use of LLMs are promising, LLM performance must be significantly improved before their deployment as decision support systems for clinical recommendations and other complex tasks.

## Introduction

Since its November 2022 launch, the Chat Generative Pre-Trained Transformer (ChatGPT; GPT-3.5-turbo) has captured widespread public attention, with media reports suggesting over 100 million monthly active users just two months after launch.^1^ Along with its successor, GPT-4, these large language models (LLMs) use a chat-based interface to respond to complex queries and solve problems.^2,3^ Although trained as general-purpose models, researchers have begun evaluating the performance of GPT-3.5-turbo and GPT-4 on clinically-relevant tasks. For instance, GPT-3.5-turbo was found to provide largely appropriate responses when asked to give simple cardiovascular disease prevention recommendations.^4^ Meanwhile, GPT-3.5-turbo responses to patients’ health questions on a public social media forum were both preferred, and rated as having higher empathy, compared to physician responses.^5^

While there are a growing number of studies that explore the uses of the GPT models across a range of clinical tasks, the majority do not use real-world clinical notes. They instead apply these models to answer questions from medical examinations such as the USMLE, solve publicly available clinical diagnostic challenges such as the *New England Journal of Medicine* (NEJM) clinicopathologic conferences, or evaluate performance on existing clinical benchmarks.^3,6–9^ This is due to the challenges associated with disclosing protected health information (PHI) with LLM providers such as OpenAI in a Health Insurance Portability and Accountability Act (HIPAA) compliant manner, where business associate agreements must be in place to allow secure processing of PHI content.^10^ This is a notable hurdle given the inherent differences between curated medical datasets, such as the USMLE question bank, and real-world clinical notes. In addition, this issue is particularly problematic when you consider that the GPT models have likely been trained on data obtained from open sources on the Internet and therefore their evaluation on existing publicly available benchmarks or tasks may be confounded by data leakage.^11^

As the availability and accessibility of these models increases, it is now critically important to better understand the potential uses and limitations of LLMs applied to actual clinical notes. In our previous work, we showed that GPT-3.5-turbo could accurately identify the higher acuity patient when provided only the clinical histories of pairs of patients presenting to the Emergency Department.^12^ This was despite a lack of additional training or fine-tuning, known as zero-shot learning.^13^ Elsewhere, Kanjee and colleagues evaluated the diagnostic ability of GPT-4 across 70 cases from the NEJM clinicopathologic conferences, obtaining a correct diagnosis in its differential in 64% of cases and as its top diagnosis in 39%.^7^ However, the ability of these general-purpose large language models to assimilate clinical information from de-identified clinical notes and return clinical recommendations is still unclear.

In this study, we sought to evaluate the zero-shot performance of GPT-3.5-turbo when prompted to provide clinical recommendations for patients evaluated in the Emergency Department. We focus on three recommendations in particular: 1) Should the patient be admitted to hospital; 2) Should the patient have radiological investigations requested; and 3) Should the patient receive antibiotics? We first evaluate performance on balanced (i.e equal numbers of positive and negative outcomes) datasets, to examine the sensitivity and specificity of GPT recommendations, before determining overall model accuracy on an unbalanced dataset that reflects real-world distributions of patients presenting to the Emergency Department.

## Results

From a total of 251,401 adult Emergency Department visits, we first created balanced samples of 10,000 ED visits for each of the three tasks (Figure 1). Using only the information provided in the *Presenting History* and *Physical Examination* sections of patients’ first ED physician note, we queried GPT-3.5-turbo to determine whether 1) the patient should be admitted to hospital, 2) the patient requires radiological investigation(s), and 3) the patient requires antibiotics, comparing its output to the ground-truth outcome extracted from the electronic health record.

**Figure 1.**
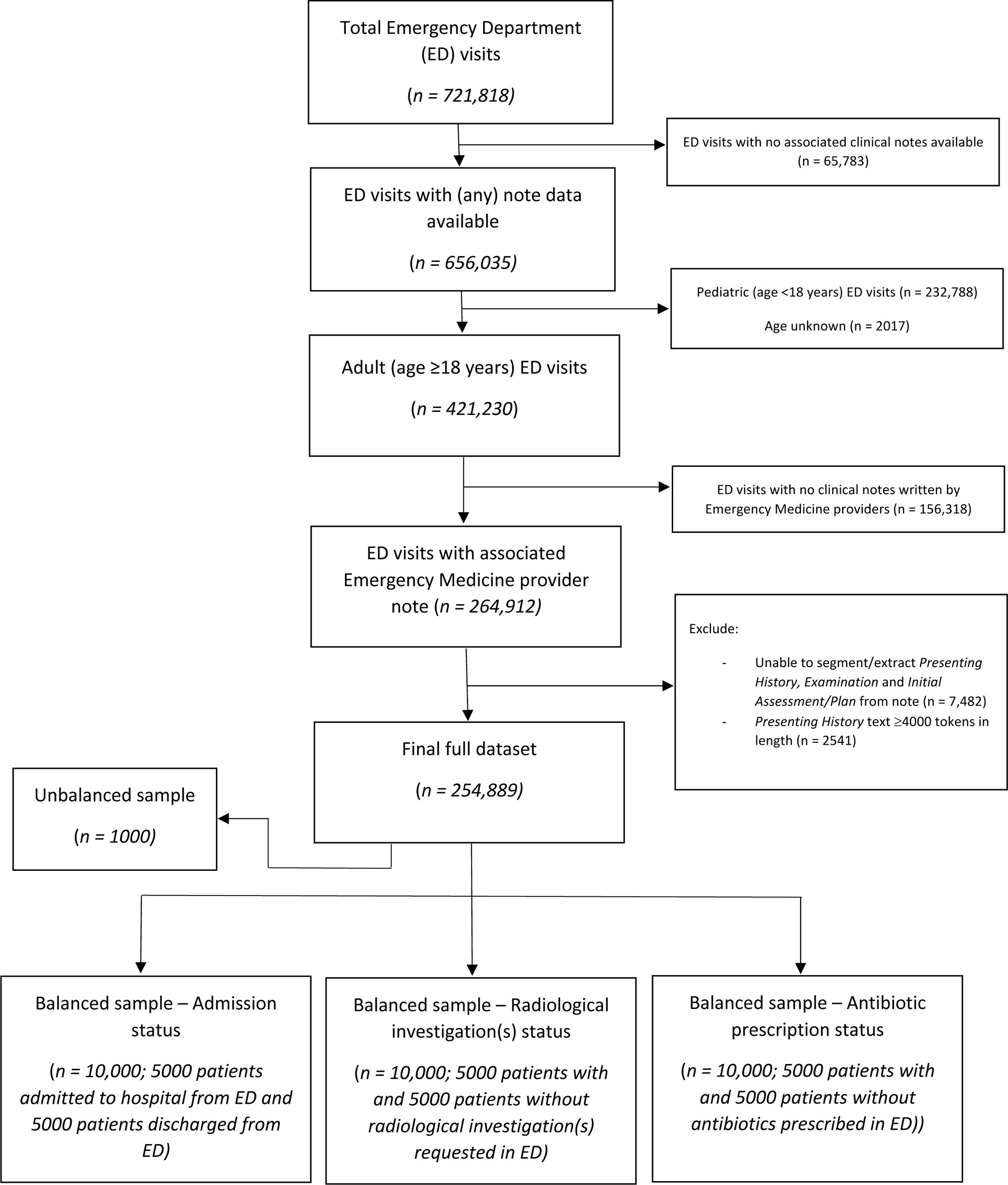
Flowchart of included Emergency Department visits and construction of both balanced (n = 10,000 samples) and unbalanced (n = 1000 sample reflecting the real-world distribution of patients presenting to the Emergency Department) datasets for the following outcomes: 1) Admission status, 2) Radiological investigation(s) status, and 3) Antibiotic prescription status.

Across all three clinical recommendation tasks, overall GPT-3.5-turbo performance was poor (Table 1). The initial prompt of ‘Please return whether the patient should be admitted to hospital / requires radiological investigation / requires antibiotics’ (Prompt A) led to high sensitivity and low specificity performance. For this prompt, GPT-3.5-turbo recommendations had a high true positive rate but similarly high false positive rate, with GPT-3.5-turbo recommending admission / radiological investigation / antibiotic prescription for the majority of cases. Altering the prompt to ‘only suggest … if absolutely required’ (Prompt B) only marginally improved specificity. The greatest performance was achieved by removing restrictions on the verbosity of GPT-3.5-turbo response (Prompt C) and adding the ‘Let’s think step by step’ chain-of-thought prompting (Prompt D). These prompts generated the highest specificity in GPT-3.5-turbo recommendations with limited effect on sensitivity.

**Table 1.**
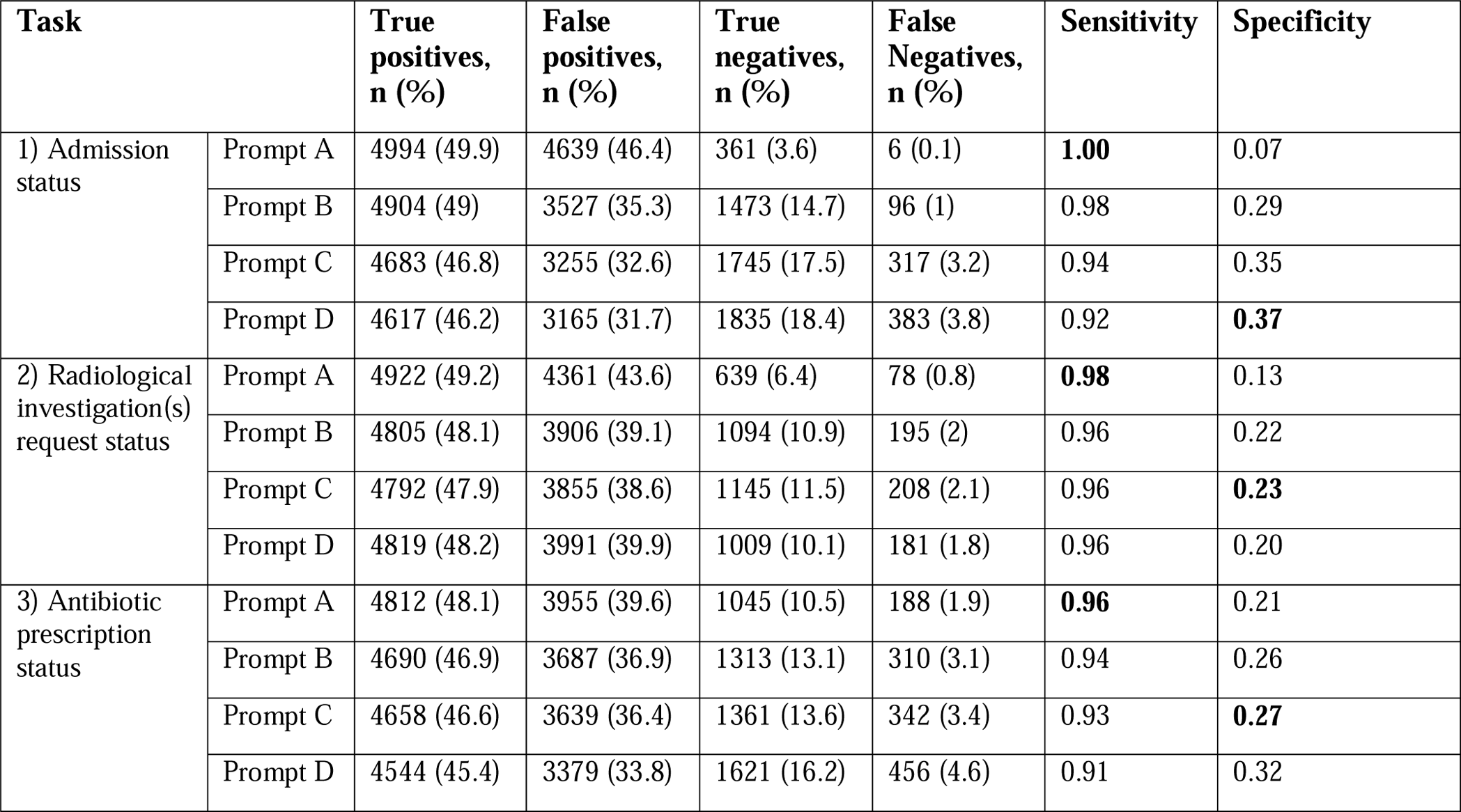
GPT-3.5-turbo performance across four iterations of prompt engineering (Prompt A-D) evaluated on a balanced n = 10,000 sample for three clinical recommendation tasks: 1) Should the patient be admitted to hospital; 2) Does the patient require radiological investigation; and 3) Does the patient require antibiotics.

| Task |  | True positives, n (%) | False positives, n (%) | True negatives, n (%) | False Negatives, n (%) | Sensitivity | Specificity |
| --- | --- | --- | --- | --- | --- | --- | --- |
| 1) Admission status | Prompt A | 4994 (49.9) | 4639 (46.4) | 361 (3.6) | 6 (0.1) | **1.00** | 0.07 |
|  | Prompt B | 4904 (49) | 3527 (35.3) | 1473 (14.7) | 96 (1) | 0.98 | 0.29 |
|  | Prompt C | 4683 (46.8) | 3255 (32.6) | 1745 (17.5) | 317 (3.2) | 0.94 | 0.35 |
|  | Prompt D | 4617 (46.2) | 3165 (31.7) | 1835 (18.4) | 383 (3.8) | 0.92 | **0.37** |
| 2) Radiological investigation(s) request status | Prompt A | 4922 (49.2) | 4361 (43.6) | 639 (6.4) | 78 (0.8) | **0.98** | 0.13 |
|  | Prompt B | 4805 (48.1) | 3906 (39.1) | 1094 (10.9) | 195 (2) | 0.96 | 0.22 |
|  | Prompt C | 4792 (47.9) | 3855 (38.6) | 1145 (11.5) | 208 (2.1) | 0.96 | **0.23** |
|  | Prompt D | 4819 (48.2) | 3991 (39.9) | 1009 (10.1) | 181 (1.8) | 0.96 | 0.20 |
| 3) Antibiotic prescription status | Prompt A | 4812 (48.1) | 3955 (39.6) | 1045 (10.5) | 188 (1.9) | **0.96** | 0.21 |
|  | Prompt B | 4690 (46.9) | 3687 (36.9) | 1313 (13.1) | 310 (3.1) | 0.94 | 0.26 |
|  | Prompt C | 4658 (46.6) | 3639 (36.4) | 1361 (13.6) | 342 (3.4) | 0.93 | **0.27** |
|  | Prompt D | 4544 (45.4) | 3379 (33.8) | 1621 (16.2) | 456 (4.6) | 0.91 | 0.32 |

To compare this performance with that of a resident physician, we took a balanced n = 200 subsample for manual annotation and compared performance between physician and machine across each of the four prompt iterations (Table 2). Notably, physician sensitivity was below that of GPT-3.5-turbo responses (0.73 vs [range: 0.93-1.00], 0.76 vs [range: 0.93-0.96] and 0.64 vs [range: 0.89-0.93] for admission, radiological investigation, and antibiotic prescription tasks, respectively), but specificity was significantly higher than GPT-3.5-turbo (0.74 vs [range: 0.07-0.40], 0.79 vs [range: 0.09-0.17] and 0.78 vs [range: 0.26-0.37]).

**Table 2.**
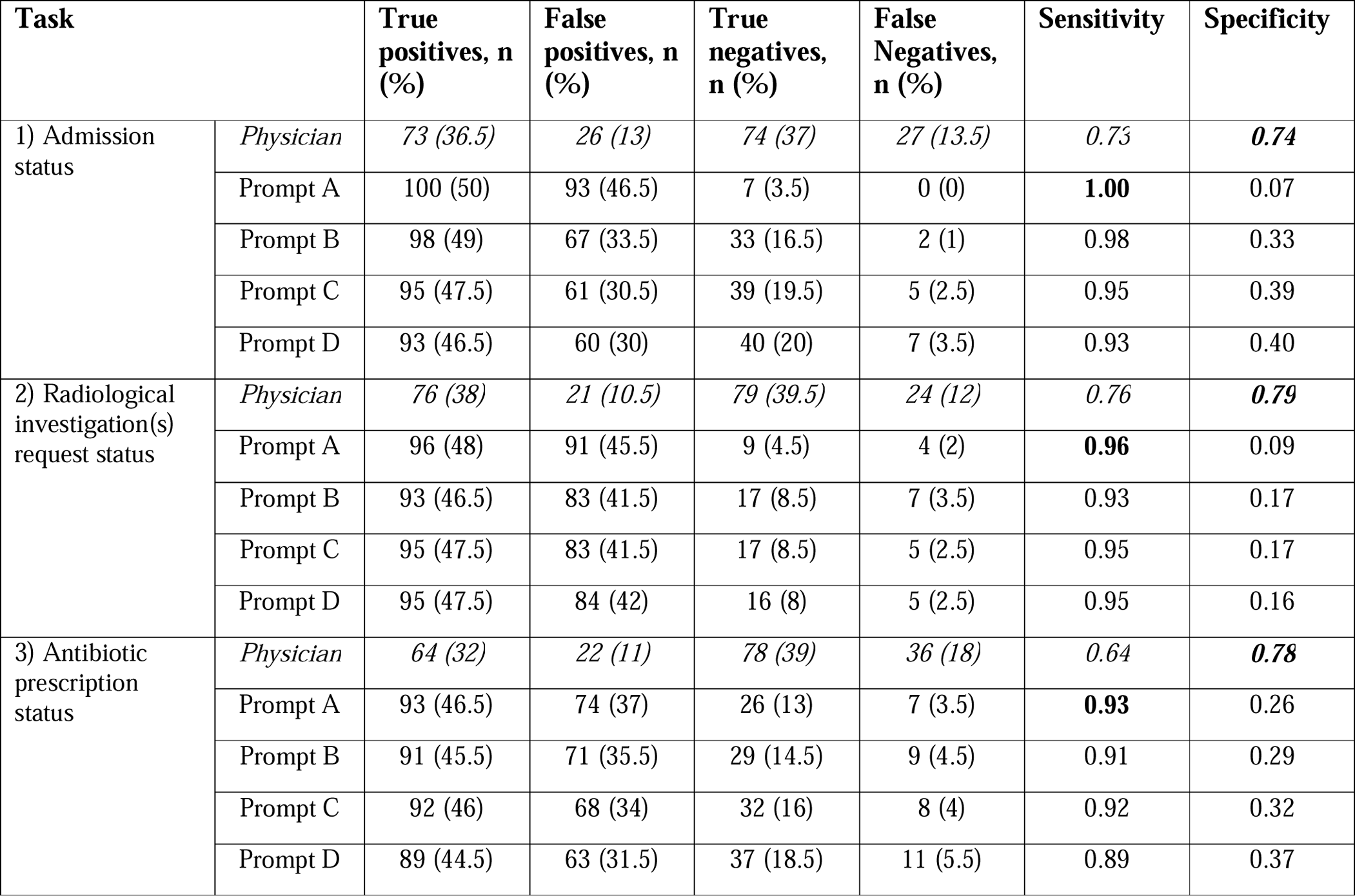
Comparison of physician and GPT-3.5-turbo performance across four iterations of prompt engineering [Prompt A-D] evaluated on a balanced n = 200 subsample for three clinical recommendation tasks: 1) Should the patient be admitted to hospital; 2) Does the patient require radiological investigation; and 3) Does the patient require antibiotics. *Physicians were provided the same prompt text as in Prompt A.

We next sought to test the performance of GPT-3.5-turbo in a more representative setting using an unbalanced, n = 1000 sample of ED visits that reflects the real-world distribution of admission, radiological investigation, and antibiotic prescription rates at our institution (Table 3). We found that the accuracy of resident physician recommendations, when evaluated against the ground-truth outcomes extracted from the electronic health record, was significantly higher than GPT-3.5-turbo recommendations: 0.83 for physician vs [range: 0.29-0.53 for GPT-3.5-turbo], 0.79 vs [range: 0.68-0.71] and 0.78 vs [range: 0.35-0.43] for admission, radiological investigation, and antibiotic prescription tasks, respectively (Figure 2; Table 3).

**Figure 2.**
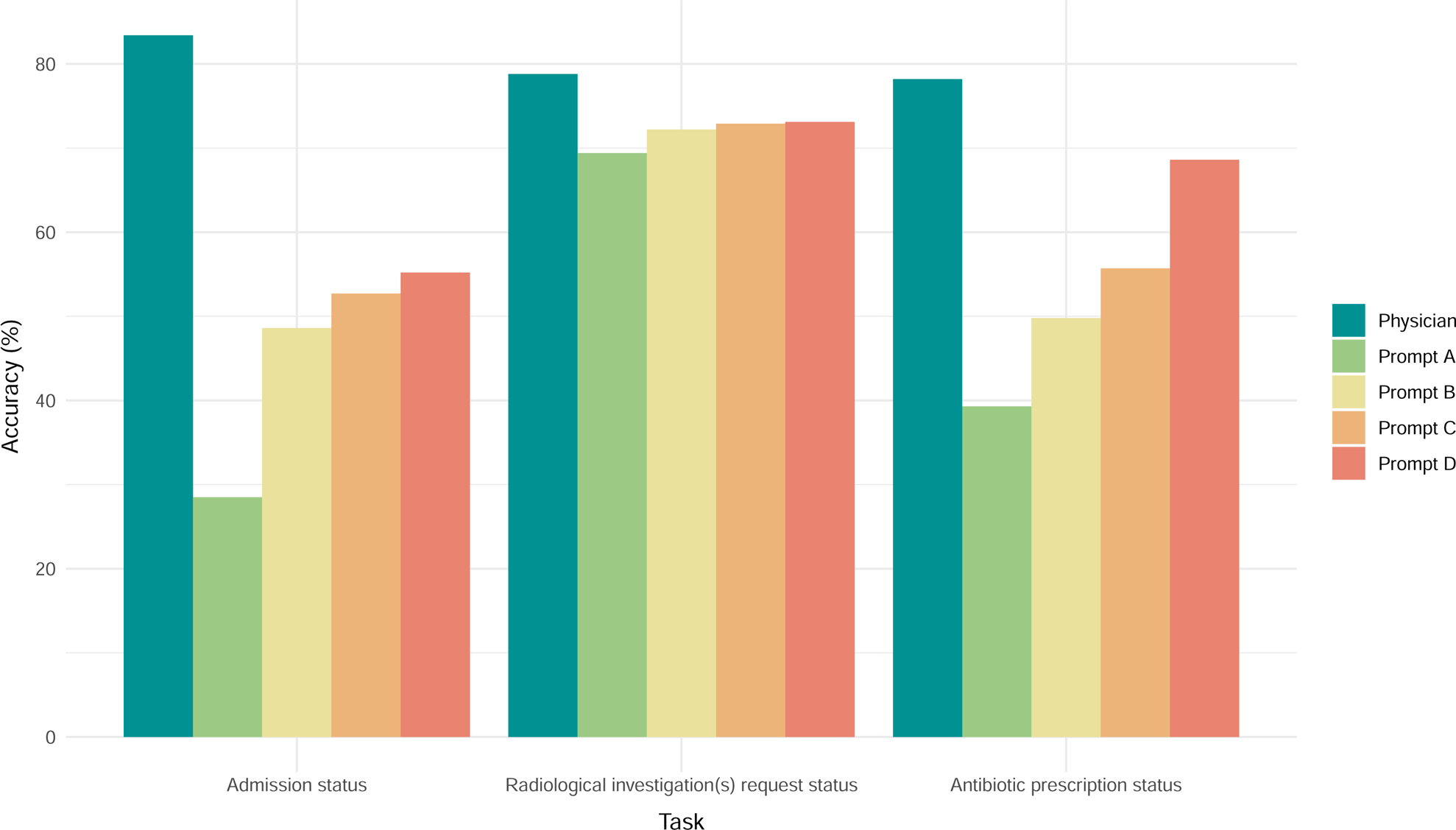
Evaluation of physician and GPT-3.5-turbo accuracy across four iterations of prompt engineering [Prompt A-D] evaluated on an unbalanced n = 1000 sample reflective of the real-world distribution of clinical recommendations among patients presenting to ED, for the following three clinical recommendation tasks: 1) Should the patient be admitted to hospital; 2) Does the patient require radiological investigation; and 3) Does the patient require antibiotics.

**Table 3.**
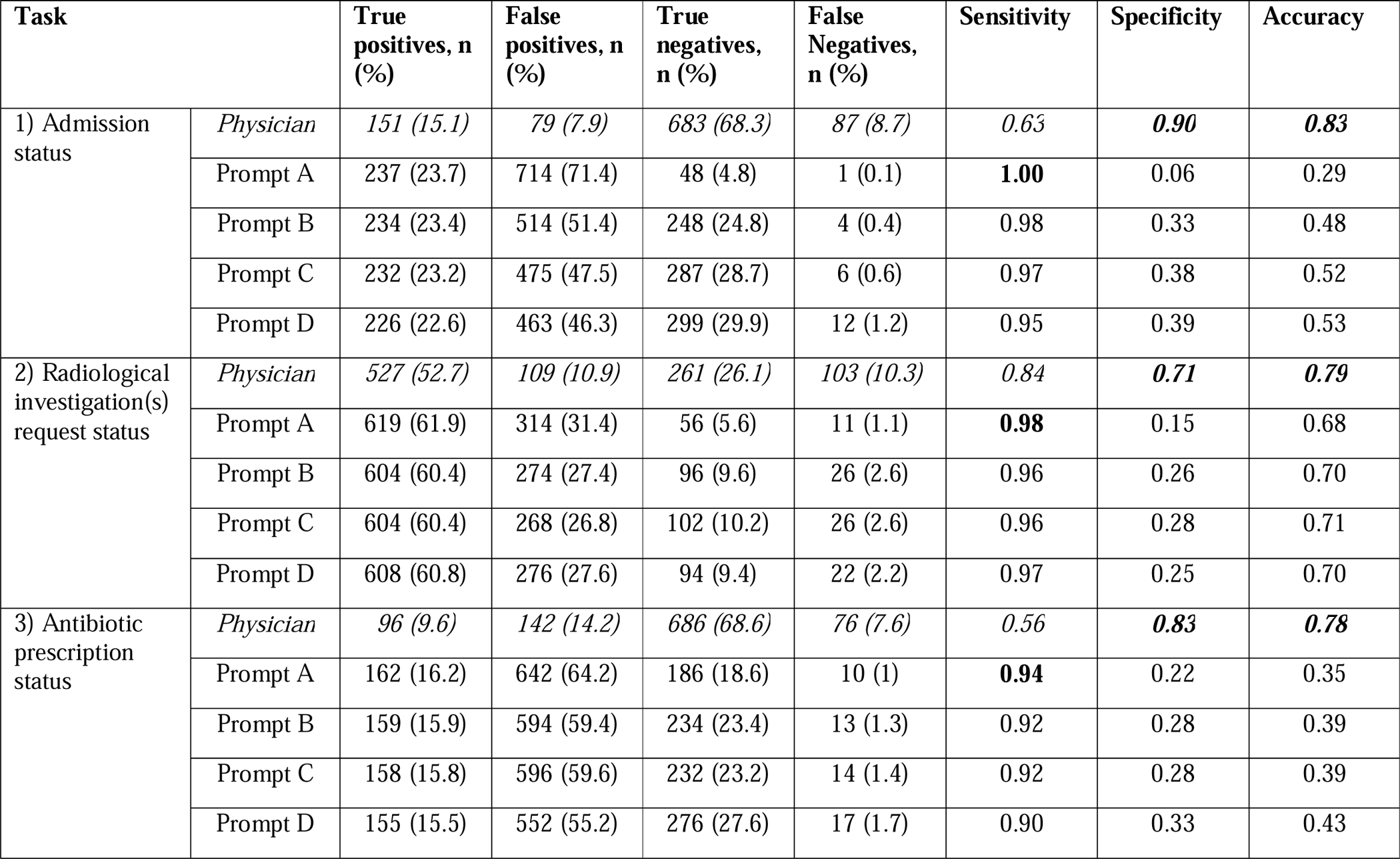
Comparison of physician and GPT-3.5-turbo performance across four iterations of prompt engineering [Prompt A-D] evaluated on an unbalanced n = 1000 sample reflective of the real-world distribution of clinical recommendations among patients presenting to ED, for the following three clinical recommendation tasks: 1) Should the patient be admitted to hospital; 2) Does the patient require radiological investigation; and 3) Does the patient require antibiotics. *Physicians were provided the same prompt text as in Prompt A.

| Task |  | True positives, n (%) | False positives, n (%) | True negatives, n (%) | False Negatives, n (%) | Sensitivity | Specificity | Accuracy |
| --- | --- | --- | --- | --- | --- | --- | --- | --- |
| 1) Admission status | Physician | 151 (15.1) | 79 (7.9) | 683 (68.3) | 87 (8.7) | 0.63 | **0.90** | **0.83** |
|  | Prompt A | 237 (23.7) | 714 (71.4) | 48 (4.8) | 1 (0.1) | **1.00** | 0.06 | 0.29 |
|  | Prompt B | 234 (23.4) | 514 (51.4) | 248 (24.8) | 4 (0.4) | 0.98 | 0.33 | 0.48 |
|  | Prompt C | 232 (23.2) | 475 (47.5) | 287 (28.7) | 6 (0.6) | 0.97 | 0.38 | 0.52 |
|  | Prompt D | 226 (22.6) | 463 (46.3) | 299 (29.9) | 12 (1.2) | 0.95 | 0.39 | 0.53 |
| 2) Radiological investigation(s) request status | Physician | 527 (52.7) | 109 (10.9) | 261 (26.1) | 103 (10.3) | 0.84 | **0.71** | **0.79** |
|  | Prompt A | 619 (61.9) | 314 (31.4) | 56 (5.6) | 11 (1.1) | **0.98** | 0.15 | 0.68 |
|  | Prompt B | 604 (60.4) | 274 (27.4) | 96 (9.6) | 26 (2.6) | 0.96 | 0.26 | 0.70 |
|  | Prompt C | 604 (60.4) | 268 (26.8) | 102 (10.2) | 26 (2.6) | 0.96 | 0.28 | 0.71 |
|  | Prompt D | 608 (60.8) | 276 (27.6) | 94 (9.4) | 22 (2.2) | 0.97 | 0.25 | 0.70 |
| 3) Antibiotic prescription status | Physician | 96 (9.6) | 142 (14.2) | 686 (68.6) | 76 (7.6) | 0.56 | **0.83** | **0.78** |
|  | Prompt A | 162 (16.2) | 642 (64.2) | 186 (18.6) | 10 (1) | **0.94** | 0.22 | 0.35 |
|  | Prompt B | 159 (15.9) | 594 (59.4) | 234 (23.4) | 13 (1.3) | 0.92 | 0.28 | 0.39 |
|  | Prompt C | 158 (15.8) | 596 (59.6) | 232 (23.2) | 14 (1.4) | 0.92 | 0.28 | 0.39 |
|  | Prompt D | 155 (15.5) | 552 (55.2) | 276 (27.6) | 17 (1.7) | 0.90 | 0.33 | 0.43 |

Lastly, in our sensitivity analyses conducted on a balanced, n = 200 subsample for each task, results were largely similar regardless of the written order of labels in the original prompt (e.g ‘0: Patient should be admitted to hospital. 1: Patient should not be admitted to hospital.’ vs ‘1: Patient should be admitted to hospital. 0: Patient should not be admitted to hospital.’) (Table S3). Reversing the order of labels in the original prompt led to almost identical results for all tasks except the antibiotic prescription task, where specificity was improved for Prompts 2-4, but at the cost of sensitivity.

## Discussion

This study represents an early, highly powered evaluation of the potential uses and limitations of GPT-3.5-turbo for generating clinical recommendations based on real-world clinical text. Across three different clinical recommendation tasks, we found that GPT-3.5-turbo performed poorly, with high sensitivity but low specificity across tasks. Model performance was marginally improved with iterations of prompt engineering, including the addition of zero-shot chain-of-thought prompting.^14^ On evaluation of an unbalanced (n = 1000) sample reflective of the real-world distribution of clinical recommendations, GPT-3.5-turbo performance was significantly worse than that of a resident physician, with 24% lower accuracy averaged across tasks.

Our results suggest that GPT-3.5-turbo is overly cautious in its clinical recommendations – it exhibits a tendency to recommend intervention for each of the three tasks and this leads to a notable number of false positive suggestions. Such a finding is problematic given the need to both prioritize hospital resource availability and reduce overall healthcare costs.^15,16^ This is also true at the patient level, where there is an increasing appreciation that excessive investigation and/or treatment may cause patients harm.^16^ It is unclear, however, what is the best balance of sensitivity/specificity to strive for amongst clinical large language models – it is likely that this balance will differ based on the particular task. The increase in GPT-3.5-turbo specificity, at the expense of sensitivity, across our iterations of prompt engineering suggests that improvements could be made bespoke to the task, though the extent to which prompt engineering alone may improve performance is unclear.

Across all three tasks, overall performance remained notably below that of a human physician. This may reflect the inherent complexity of clinical decision making, where clinical recommendations may be influenced not only by the patient’s intrinsic clinical status, but also by patient preference, current resource availability and other external factors.

Before large language models can be integrated within the clinical environment, it is important to fully understand both their capabilities and limitations. Otherwise, there is a risk of unintended harmful consequences, especially if models have been deployed at scale.^17,18^ Current research deploying large language models, particularly the current state-of-the-art GPT models, on real-world clinical text is limited. Recent work from our group has demonstrated accurate performance of GPT-3.5-turbo in both assessing patient clinical acuity in the Emergency Department and extracting detailed oncologic history and treatment plans from medical oncology notes.^19^ Elsewhere, GPT-3.5-turbo has been used to convert radiology reports into plain language, to classify whether statements of clinical recommendations in scientific literature constitute health advice, and to accurately classify five diseases from discharge summaries in the MIMIC-III dataset.^20–22^ Much of the current literature focuses on the strengths of large language models such as GPT-3.5-turbo and GPT-4.^3,9,12,19^ However, it is equally important to identify areas of medicine in which LLMs do not perform well. For example, in one evaluation of GPT-4’s ability to diagnose dementia from a set of structured features, GPT-4 did not surpass the performance of traditional AI tools, while fewer than 20% of GPT-3.5-turbo and GPT-4 responses submitted to a clinical informatics consult service were found to be concordant with existing reports.^23,24^ While early signs of the utility of large language models in medicine are promising, our findings suggest that there remains significant room for improvement, especially in more challenging tasks such as complex clinical decision making.

This study has several limitations. Firstly, it is possible that, for each task, not all the information which led to the real-life clinical recommendation extracted from the electronic health record was present in the *Presenting History* and *Physical Examination* sections of the ED physician note. For instance, radiological investigations requested following the Emergency Medicine physician review may lead to unexpected and/or incidental findings which were not detected during the initial review and may warrant admission or antibiotic prescription. However, even with this limitation, physician classification performance remained a very respectable 78-83% accuracy across the three tasks, suggesting it is challenging, but not impossible, to make accurate clinical recommendations based on the available clinical text. Secondly, we only trialled three iterations of prompt engineering, in addition to our initial prompt, and this was done in a zero-shot manner. Further attempts to refine the provided prompt, or incorporate few-shot examples for in-context learning, may improve model performance.^13,25–27^ Lastly, this study did not evaluate the performance of the recently released, more advanced GPT-4 model. It is possible that GPT-4 performance may surpass that of GPT-3.5-turbo in these more complex reasoning tasks, though the ability to test this at a similar scale is limited by the increased costs associated with GPT-4 usage across a sample of this size. Similarly, evaluation of the performance of other natural language processing models, such as a fine-tuned BioClinicalBERT model or bag-of-word-based and other simpler techniques, has not been performed.^28^ It is possible that these more traditional NLP models, which are typically trained or fine-tuned on a large training set of data, may outperform the zero-shot performance of GPT-like large language models.^21^

## Methods

The UCSF Information Commons contains deidentified structured clinical data as well as deidentified clinical text notes, deidentified and externally certified as previously described.^29^ The UCSF Institutional Review Board determined that this use of the deidentified data within the UCSF Information Commons is not human participants research and therefore was exempt from further approval and informed consent.

We identified all adult visits to the University of California San Francisco (UCSF) Emergency Department (ED) from 2012 to 2023 with an ED Physician note present within Information Commons (Figure 1). Regular expressions were used to extract the *Presenting History* (consisting of ‘Chief Complaint’, ‘History of Presenting Illness’ and ‘Review of Systems’) and *Physical Examination* sections from each note (Supplementary File 1).

We sought to evaluate GPT-3.5-turbo performance on three binary clinical recommendation tasks, corresponding to the following outcomes: 1) Admission status – whether the patient should be admitted from ED to hospital. 2) Radiological investigation(s) request status – whether an X-ray, US scan, CT scan, or MRI scan should be requested during the ED visit. 3) Antibiotic prescription status – whether antibiotics should be ordered during the ED visit.

For each of the three outcomes, we randomly selected a balanced sample of 10,000 ED visits to evaluate GPT-3.5-turbo performance (Figure 1). Using its secure, HIPAA-compliant Application Programming Interface (API) through Microsoft Azure, we provided GPT-3.5-turbo (model *gpt-3.5-turbo-0301*) the *Presenting History* and *Physical Examination* sections of the ED Physician’s note for each ED visit and queried it to determine if 1) the patient should be admitted to hospital, 2) the patient requires radiological investigation, and 3) the patient should be prescribed antibiotics. GPT-3.5-turbo performance was evaluated against the ground-truth outcome extracted from the electronic health record. Separately, a resident blinded to both the GPT-3.5-turbo labels and ground-truth labels reviewed a balanced n = 200 subsample for each of the three tasks to allow a comparison of human and machine performance. The following evaluation metrics were calculated: true positive rate, true negative rate, false positive rate, false negative rate, sensitivity and specificity.

We subsequently experimented with three iterations of prompt engineering (Table S1, Supplementary File 1) to test if modifications to the initial prompt could improve GPT-3.5-turbo performance. Chain-of-thought (CoT) prompting is a method found to improve the ability of large language models to perform complex reasoning by decomposing multi-step problems into a series of intermediate steps.^25^ This can be done in a zero-shot manner (zero-shot-CoT), with large language models shown to be decent zero-shot reasoners by adding a simple prompt, ‘Let’s think step by step’ to facilitate step-by-step reasoning before answering each question.^14^ Alternatively, few-shot chain-of-thought prompting can be used, with additional examples of prompt and answer pairs either manually (manual CoT) or computationally (e.g auto-CoT) provided and concatenated with the prompt of interest.^25,26^ Current understanding of the impact of zero-shot-CoT, manual CoT, and auto-CoT prompt engineering techniques applied to clinical text is limited. In this work, we sought to focus on zero-shot-CoT and investigate the effect of adding ‘Let’s think step by step’ to the prompt on model performance.

Our *initial prompt* (Prompt A) simply asked GPT-3.5-turbo to return whether the patient should be e.g. admitted to hospital, without any additional explanation. We additionally attempted to engineer prompts to a) reduce the high false positive rate of GPT-3.5-turbo recommendations (Prompt B) and b) examine whether zero-shot chain-of-thought prompting could improve GPT-3.5-turbo performance (Prompts C and D). Attempting to reduce the high GPT-3.5-turbo false positive rate, Prompt B was constructed by adding an additional sentence to Prompt A: ‘Only suggest \**clinical recommendation\** if absolutely required’. This modification was kept for Prompts C and D, which were constructed to examine chain-of-thought prompting. Because chain-of-thought prompting is most effective when the LLM provides reasoning in its output, we removed the instruction ‘Please do not return any additional explanation’ from Prompts C and D, and added the chain-of-thought prompt ‘Let’s think step by step’ to Prompt D, increasing GPT-3.5-turbo response verbosity (Table S2, Supplementary File 1). Prompt C therefore served as a baseline for comparison of GPT-3.5-turbo performance when it is permitted to return additional explanation (in addition to its outcome recommendation), allowing comparisons with both Prompt A (where no additional explanations were allowed in the prompt) and Prompt D (where the effect of chain-of-thought prompting was examined).

To evaluate the performance of GPT-3.5-turbo in a real-world setting, we constructed a random, unbalanced sample of 1000 ED visits where the distribution of patient outcomes (i.e. admission status, radiological investigation(s) request status and antibiotic prescription status) mirrored the distributions of patients presenting to ED from our main cohort. The *Presenting History* and *Physical Examination* sections of the ED Physician’s note for each ED visit were again passed to the GPT-3.5-turbo API in an identical manner to the balanced datasets, while a resident physician manually labelled the entire sample to allow human vs machine comparison. Classification accuracy was calculated in addition to the aforementioned evaluation metrics utilised for the balanced datasets to provide a summative evaluation metric for this real-world simulated task.

### Sensitivity analysis

Due to the stochastic nature of large language models, it is possible that the order of labels reported in the original prompt may affect the subsequent labels returned. To test this, we conducted a sensitivity analysis on a balanced n = 200 subsample for each outcome where the positive outcome was referenced before the negative outcome in the initial prompt (e.g. ‘1: Patient should be admitted to hospital’ precedes ‘0: Patient should not be admitted to hospital’ in the GPT-3.5-turbo prompt).

## Acknowledgements

The authors acknowledge the use of the UCSF Information Commons computational research platform, developed and supported by UCSF Bakar Computational Health Sciences Institute. The authors also thank the UCSF AI Tiger Team, Academic Research Services, Research Information Technology, and the Chancellor’s Task Force for Generative AI for their software development, analytical and technical support related to the use of Versa API gateway (the UCSF secure implementation of large language models and generative AI via API gateway), Versa chat (the chat user interface), and related data asset and services.

## Data availability

The code accompanying this manuscript is available at https://github.com/cykwilliams/GPT-3.5-Clinical-Recommendations-in-Emergency-Department/.

