## Supplemental File 1 for "Evaluating the use of GPT-3.5-turbo to provide clinical recommendations in the Emergency Department"

*Cohort selection*

Only adult patient (≥18 years) Emergency Department (ED) visits were considered in this study. ED visits with no associated clinical notes were excluded, as were visits with clinical notes written only by non-Emergency Medicine providers. If more than one Emergency Medicine provider note was available for a particular ED visit, the earliest note was selected. In the case of multiple notes with the same chart time, the longest note (by word count) was selected.

*Note pre-processing & segmentation*

Clinical notes were minimally preprocessed - only new lines and extra spaces were removed. A series of Regular Expressions were used to examine the structure of notes, confirming the presence/absence of the following note headers: ‘Chief Complaint’; ‘Review of Systems’; ‘Physical Exam’; ‘Initial Assessment’ and/or ‘ED Course’. For each clinical note, we extracted all text from:

1) Clinical History: section ‘Chief Complaint’ (inclusive) to ‘Physical Exam’, representing the full history of each patient’s ED visit, including both their Presenting Complaint/History of Presenting Complaint and Systems Review;

2) Examination: section ‘Physical Exam’ (inclusive) to either ‘Initial Assessment’ or ‘ED course’, representing the Physical Examination findings; and

3) Assessment/Plan: from ‘Initial Assessment’ or ‘ED course’ to note end, representing the clinician’s Impression/Assessment and Plan.

*Tokenisation*

A sample of the segmented note text was examined to confirm proper extraction. Only ED visits in which all three sections of the accompanying Emergency Medicine Provider note could be segmented and extracted were included. For this study, only text from the Clinical History and Examination sections of patients’ clinical notes was analysed by GPT-3.5-turbo.

The number of tokens for each section was calculated using the tiktoken tokenizer module recommended by Open AI. Tokens can be thought of as pieces of words which form the input of large language models; 100 tokens are approximately equal to 75 words. Notably, GPT-3.5-turbo has a maximum limit of 4096 tokens shared between prompt (input) and completion (output). Consequently, we filtered our dataset to exclude the 2541 ED visits with a note ≥4000 tokens in length (Figure 1).

*GPT3.5-turbo Prompt Engineering*

We used GPT-3.5-turbo to perform zero shot classification of whether patients should 1) be admitted, 2) receive radiological investigation(s) and 3) be prescribed antibiotics. We deployed the following text for prompting GPT-3.5-turbo:

| **Initial prompt** | |
| --- | --- |
| Admission status | “You are an Emergency Department physician. Below are the symptoms of a patient presenting to the Emergency Department. Please return whether the patient should be admitted to hospital. Please return one of two answers: '0: Patient should not be admitted to hospital' '1: Patient should be admitted to hospital' Please do not return any additional explanation.” |
| Radiological investigation(s) status | “You are an Emergency Department physician. Below are the symptoms of a patient presenting to the Emergency Department. Please return whether the patient requires radiological investigation (e.g X-ray, ultrasound scan, CT scan or MRI scan). Please return one of two answers: '0: Patient does not require radiological investigation' '1: Patient requires radiological investigation' Please do not return any additional explanation." |
| Antibiotic prescription status | “You are an Emergency Department physician. Below are the symptoms and clinical examination findings of a patient presenting to the Emergency Department. Please return whether the patient requires antibiotics. Please return one of two answers: '0: Patient does not require antibiotics' '1: Patient requires antibiotics'. Please do not return any additional explanation.” |
| **Only recommend if absolutely required** | |
| Admission status | “You are an Emergency Department physician. Below are the symptoms of a patient presenting to the Emergency Department. Please return whether the patient should be admitted to hospital. **Only suggest admission to hospital if absolutely required.** Please return one of two answers: '0: Patient should not be admitted to hospital' '1: Patient should be admitted to hospital' Please do not return any additional explanation." |
| Radiological investigation(s) status | "You are an Emergency Department physician. Below are the symptoms of a patient presenting to the Emergency Department. Please return whether the patient requires radiological investigation (e.g X-ray, ultrasound scan, CT scan or MRI scan). **Only suggest radiological investigation if absolutely required.** Please return one of two answers: '0: Patient does not require radiological investigation' '1: Patient requires radiological investigation' Please do not return any additional explanation." |
| Antibiotic prescription status | “You are an Emergency Department physician. Below are the symptoms and clinical examination findings of a patient presenting to the Emergency Department. Please return whether the patient requires antibiotics. **Only suggest antibiotics if absolutely required**. Please return one of two answers: '0: Patient does not require antibiotics' '1: Patient requires antibiotics'. Please do not return any additional explanation.” |
| **Chain-of-thought prompting (baseline)** | |
| Admission status | “You are an Emergency Department physician. Below are the symptoms of a patient presenting to the Emergency Department. Please return whether the patient should be admitted to hospital. Only suggest admission to hospital if absolutely required. Please return one of two answers: '0: Patient should not be admitted to hospital' '1: Patient should be admitted to hospital' ~~Please do not return any additional explanation.~~" |
| Radiological investigation(s) status | "You are an Emergency Department physician. Below are the symptoms of a patient presenting to the Emergency Department. Please return whether the patient requires radiological investigation (e.g X-ray, ultrasound scan, CT scan or MRI scan). Only suggest radiological investigation if absolutely required. Please return one of two answers: '0: Patient does not require radiological investigation' '1: Patient requires radiological investigation' ~~Please do not return any additional explanation.~~" |
| Antibiotic prescription status | “You are an Emergency Department physician. Below are the symptoms and clinical examination findings of a patient presenting to the Emergency Department. Please return whether the patient requires antibiotics. Only suggest antibiotics if absolutely required. Please return one of two answers: '0: Patient does not require antibiotics' '1: Patient requires antibiotics'. ~~Please do not return any additional explanation.~~” |
| **Chain-of-thought prompting – ‘Let’s think step by step’** | |
| Admission status | “You are an Emergency Department physician. Below are the symptoms of a patient presenting to the Emergency Department. Please return whether the patient should be admitted to hospital. **Let's think step by step.** Only suggest admission to hospital if absolutely required. Please return one of two answers: '0: Patient should not be admitted to hospital' '1: Patient should be admitted to hospital'" |
| Radiological investigation(s) status | "You are an Emergency Department physician. Below are the symptoms of a patient presenting to the Emergency Department. Please return whether the patient requires radiological investigation (e.g X-ray, ultrasound scan, CT scan or MRI scan). **Let's think step by step.** Only suggest radiological investigation if absolutely required. Please return one of two answers: '0: Patient does not require radiological investigation' '1: Patient requires radiological investigation'" |
| Antibiotic prescription status | “You are an Emergency Department physician. Below are the symptoms and clinical examination findings of a patient presenting to the Emergency Department. Please return whether the patient requires antibiotics. **Let's think step by step.** Only suggest antibiotics if absolutely required. Please return one of two answers: '0: Patient does not require antibiotics' '1: Patient requires antibiotics'. |

*GPT-3.5-turbo and human evaluation*

Prompts were sent to the GPT-3.5-turbo Application Programming Interface (API) (model = ‘gpt-3.5-turbo-0301’, role = ‘user’, temperature = 0; all other settings at default values) via the HIPAA-compliant, UCSF Secure Azure OpenAI environment and responses from the API were recorded.

Separately, a resident physician blinded to both the GPT-3.5-turbo labels and ground-truth labels reviewed the Clinical History and Examination sections of both 1) n = 200 subsamples of each of the three balanced datasets and 2) the entire unbalanced, n = 1000 dataset to provide human recommendations in each of the three tasks.

| **Task** | **Prompt** | **Word count, mean (sd)** |
| --- | --- | --- |
| Admission status | A | 7.0 (0.2) |
|  | B | 7.2 (0.5) |
|  | C | 47.6 (26.5) |
|  | D | 54.5 (27.3) |
| Radiological investigation(s) request status | A | 17.7 (8.4) |
|  | B | 18.5 (8.8) |
|  | C | 24.0 (13.6) |
|  | D | 26.3 (16.1) |
| Antibiotic prescription status | A | 4.6 (2.5) |
|  | B | 5.0 (3.9) |
|  | C | 19.6 (18.6) |
|  | D | 31.7 (24.4) |

Table S2. Word counts of GPT-3.5-turbo response text for different iterations of prompt engineering [Prompt A-D] evaluated on the balanced n = 10000 sample for three clinical recommendation tasks: 1) Should the patient be admitted to hospital; 2) Does the patient require radiological investigation; and 3) Does the patient require antibiotics. Abbreviations: sd = standard deviation.

| **Model** | **Task** | | **True positives, n (%)** | **False positives, n (%)** | **True negatives, n (%)** | **False Negatives, n (%)** | **Sensitivity** | **Specificity** |
| --- | --- | --- | --- | --- | --- | --- | --- | --- |
| a) GPT-3.5-turbo | Admission status | *Physician* | *73 (36.5)* | *26 (13)* | *74 (37)* | *27 (13.5)* | *0.73* | ***0.74*** |
|  |  | Prompt A | 100 (50) | 93 (46.5) | 7 (3.5) | 0 (0) | **1.00** | 0.07 |
|  |  | Prompt B | 98 (49) | 67 (33.5) | 33 (16.5) | 2 (1) | 0.98 | 0.33 |
|  |  | Prompt C | 95 (47.5) | 61 (30.5) | 39 (19.5) | 5 (2.5) | 0.95 | 0.39 |
|  |  | Prompt D | 93 (46.5) | 60 (30) | 40 (20) | 7 (3.5) | 0.93 | 0.40 |
|  | Radiological investigation(s) request status | *Physician* | *76 (38)* | *21 (10.5)* | *79 (39.5)* | *24 (12)* | *0.76* | ***0.79*** |
|  |  | Prompt A | 96 (48) | 91 (45.5) | 9 (4.5) | 4 (2) | **0.96** | 0.09 |
|  |  | Prompt B | 93 (46.5) | 83 (41.5) | 17 (8.5) | 7 (3.5) | 0.93 | 0.17 |
|  |  | Prompt C | 95 (47.5) | 83 (41.5) | 17 (8.5) | 5 (2.5) | 0.95 | 0.17 |
|  |  | Prompt D | 95 (47.5) | 84 (42) | 16 (8) | 5 (2.5) | 0.95 | 0.16 |
|  | Antibiotic prescription status | *Physician* | *64 (32)* | *22 (11)* | *78 (39)* | *36 (18)* | *0.64* | ***0.78*** |
|  |  | Prompt A | 93 (46.5) | 74 (37) | 26 (13) | 7 (3.5) | **0.93** | 0.26 |
|  |  | Prompt B | 91 (45.5) | 71 (35.5) | 29 (14.5) | 9 (4.5) | 0.91 | 0.29 |
|  |  | Prompt C | 92 (46) | 68 (34) | 32 (16) | 8 (4) | 0.92 | 0.32 |
|  |  | Prompt D | 89 (44.5) | 63 (31.5) | 37 (18.5) | 11 (5.5) | 0.89 | 0.37 |
| b) GPT-3.5-turbo (reversed prompt) | Admission status | *Physician* | *73 (36.5)* | *26 (13)* | *74 (37)* | *27 (13.5)* | *0.73* | ***0.74*** |
|  |  | Prompt A | 100 (50) | 93 (46.5) | 7 (3.5) | 0 (0) | **1.00** | 0.07 |
|  |  | Prompt B | 98 (49) | 65 (32.5) | 35 (17.5) | 2 (1) | 0.98 | 0.35 |
|  |  | Prompt C | 96 (48) | 59 (29.5) | 41 (20.5) | 4 (2) | 0.96 | 0.41 |
|  |  | Prompt D | 92 (46) | 54 (27) | 46 (23) | 8 (4) | 0.92 | 0.46 |
|  | Radiological investigation(s) request status | *Physician* | *76 (38)* | *21 (10.5)* | *79 (39.5)* | *24 (12)* | *0.76* | ***0.79*** |
|  |  | Prompt A | 94 (47) | 85 (42.5) | 15 (7.5) | 6 (3) | **0.94** | 0.15 |
|  |  | Prompt B | 90 (45) | 73 (36.5) | 27 (13.5) | 10 (5) | 0.90 | 0.27 |
|  |  | Prompt C | 90 (45) | 69 (34.5) | 31 (15.5) | 10 (5) | 0.90 | 0.31 |
|  |  | Prompt D | 89 (44.5) | 70 (35) | 30 (15) | 11 (5.5) | 0.89 | 0.30 |
|  | Antibiotic prescription status | *Physician* | *64 (32)* | *22 (11)* | *78 (39)* | *36 (18)* | *0.64* | ***0.78*** |
|  |  | Prompt A | 94 (47) | 72 (36) | 28 (14) | 6 (3) | **0.94** | 0.28 |
|  |  | Prompt B | 81 (40.5) | 56 (28) | 44 (22) | 19 (9.5) | 0.81 | 0.44 |
|  |  | Prompt C | 76 (38) | 48 (24) | 52 (26) | 24 (12) | 0.76 | 0.52 |
|  |  | Prompt D | 63 (31.5) | 33 (16.5) | 67 (33.5) | 37 (18.5) | 0.63 | 0.67 |

Table S3. Sensitivity analysis: comparison of physician and a) GPT-3.5-turbo performance, b) GPT-3.5-turbo performance with reversed prompt across four iterations of prompt engineering [Prompt A-D] evaluated on a balanced n = 200 subsample for three clinical recommendation tasks: 1) Should the patient be admitted to hospital; 2) Does the patient require radiological investigation; and 3) Does the patient require antibiotics.
